# Women’s empowerment and life stage: assessing intersectional differences in contraceptive method mix in sub-Saharan Africa

**DOI:** 10.1101/2025.02.14.25322228

**Authors:** Franciele Hellwig, Yohannes Dibaba Wado, Cheikh M Faye, Jennifer Requejo, Leontine Alkema, Rornald Muhumuza Kananura, Ties Boerma, Aluísio JD Barros

## Abstract

**Background:** Women’s empowerment positively impacts family planning in sub-Saharan Africa, but little is known about its relationship with contraceptive method mix. Considering that contraceptive needs vary across life stages, we aimed to explore the intersectional differences in contraceptive method mix according to age and level of empowerment in sub-Saharan Africa.

**Methods:** We analyzed data from Demographic and Health Surveys conducted from 2015 to 2022 across 28 countries in West & Central and Eastern & Southern Africa. Within each region, we calculated pooled estimates of demand for family planning satisfied by any method (DFPS) and contraceptive method mix, stratifying by women’s age and empowerment level according to the SWPER Global index.

**Results:** Our sample included 138,374 married women of reproductive age. Across both regions, DFPS increased with empowerment, especially among women older than 19. Among adolescents, a substantial increase was only identified in terms of decision-making. Contraceptive method mix was skewed towards injectables and implants, but their shares dropped as empowerment increased. This trend appeared across all age groups and empowerment domains, with the largest declines observed for social independence. High-empowered women had a more diversified method mix, with adolescents and young women showing increased use of condoms, pills, and fertility-awareness methods, while women over 34 relied more on IUDs and sterilization.

**Conclusion:** Our findings reveal that age, empowerment, and contraceptive method mix intersect in diverse ways. Tailoring family planning policies and counseling while addressing empowerment drivers could enhance informed contraceptive choices and better meet women’s reproductive health goals.

## Introduction

Expanding the use of contraception is a key commitment in several international initiatives, such as the Sustainable Development Goals (SDGs), the Global Strategy for Women’s, Children’s, and Adolescents’ Health (2016-2030), and the Family Planning 2030 (FP2030). In response to these initiatives, massive investments have been made to promote family planning in low- and middle-income countries (1). While these investments have significantly increased contraceptive use worldwide, less attention has been given to the quality of family planning services being provided.

Family planning is defined as the individual’s right to decide the number and spacing of their children and to choose the means by which this can be achieved (2,3). Today, when a variety of contraceptive methods are available, women’s method choice should ideally reflect their needs and preferences (2). However, while this choice is directly influenced by women’s and couples’ age, parity, desired family size, known side effects, and health conditions, it is also influenced by a complex interaction between these individual aspects with socioeconomic, cultural, structural, and political factors.

In sub-Saharan Africa, family planning efforts are relatively recent compared to other regions and, as a result, have been occurring in a context in which a wide range of contraceptive methods is already available (4). However, the method mix in the region is still highly skewed though changes were identified over the last few decades (5). Family planning strategies adopted in the region have predominantly focused on injectables and implants, which can be easily provided even by community health workers, offer longer-term protection, and, once inserted, are not user-controlled (6).

It is expected that family planning programs, particularly in areas with weak health systems, concentrate on one or two methods to increase and maintain coverage. However, once a method becomes culturally established, it tends to be favored by both healthcare providers and users, leading to a form of path dependency (7,8). Evidence shows that healthcare providers sometimes use various strategies to promote certain methods, ranging from enthusiasm for a particular method to practices like falsifying information to encourage adoption (8,9).

There is substantial evidence showing the positive impact of women’s empowerment on women’s practice of family planning (10–13). More empowered women are more capable of overcoming sociocultural norms to adopt family planning practices and access contraception (10,13). However, the magnitude of this effect varies significantly across sub-Saharan African countries, with countries in West & Central Africa presenting lower levels of women’s empowerment and access to family planning compared to those in Eastern & Southern Africa (14–17). Additionally, the influence of empowerment varies across different life stages, as age affects both reproductive needs and the sociocultural pressures women face. The interaction between age, empowerment, and the type of contraceptive used has not yet been explored in the sub-Saharan African context. Understanding how empowerment shapes contraceptive choices at different life stages can help design age-appropriate interventions that better meet the reproductive needs of women. Using the SWPER Global (15), a standard measure of women’s empowerment based on ever-married/partnered women, this study aims to address this gap by analyzing how the intersection of women’s empowerment and age shapes the contraceptive method mix in West & Central Africa and Eastern & Southern Africa, providing evidence to guide more targeted family planning strategies in the regions.

## Methods

### Data

We used publicly available data from the most recent Demographic and Health Surveys (DHS) carried out since 2015 in sub-Saharan Africa countries. These are publicly available, nationally representative cross-sectional surveys with information on women of reproductive age (15-49 years). A total of 28 surveys were included in the analyses, representing countries with data on family planning and women’s empowerment for ever-married women or those in union.

### Outcomes

We first explored demand for family planning satisfied (DFPS), defined as the proportion of women in need of contraception who were using (or whose partner was using) any contraceptive method. A woman was considered in need of contraception if she was fecund and did not want to become pregnant within the next 2 years, or if she was unsure about whether or when she wanted to become pregnant. Pregnant women with a mistimed or unintended pregnancy are also considered in need of contraception (18).

To study the method mix, we calculated the proportion of users of each of the following contraceptive methods. Modern contraceptives, defined as medical procedures or technological products (19), which include pills, injectables, male condoms, intrauterine devices (IUDs), implants, sterilization (male or female), or other modern. And non-modern methods: lactational amenorrhea (LAM), withdrawal, fertility-awareness based methods (including periodic abstinence, basal body temperature, TwoDay Method, and Standard Days Method), and other non-modern methods.

### Stratifiers

Women’s empowerment was measured using the SWPER Global (15), an individual-level indicator estimated for married women of reproductive age. The SWPER was derived using principal component analyses based on 14 DHS questions and is comprised of three domains of women’s empowerment: (1) social independence, related to access to information, education and age of marriage and first birth; (2) decision-making, related to making decisions on important household matters and (3) attitude towards violence, related to how much the woman rejects domestic violence against the wife. The resulting scores were standardized so that positive values represent above-average levels of empowerment across LMICs used to derive the indicator, while negative values represent the opposite. The zero value represents average empowerment across the LMICs used to derive the indicator. Based on the continuous SWPER scores and standardized cut-offs, each domain was categorized into low, medium, and high levels of empowerment (15). Women’s age at the time of the survey was categorized into three groups (15-19, 20-34, and 35-49 years). All results were stratified and presented according to the UNICEF classification: West & Central Africa and Eastern & Southern Africa.

Exploratory analyses were conducted to ensure comparability in the empowerment levels across the three empowerment categories, age groups, and regions. Additionally, to ensure that women classified into low, medium, or high empowerment levels had comparable levels of empowerment across key contextual factors, a sensitivity analysis was conducted comparing the mean empowerment scores of the SWPER Global across wealth quintiles and urban-rural residence within each category of empowerment level and age group, stratified by region.

### Statistical analyses

Descriptive analyses were conducted with the countries pooled together. Data from the 28 countries were aggregated with weighting for population size in the median year (2019).

All analyses were performed using Stata software version 18.0 (StataCorp LLC, College Station, TX) and adjusted for the sample design, including sample weights, clusters, and strata. All analyses relied on publicly available anonymized databases. Institutions and national agencies in each country obtained ethics approval for the surveys.

## Results

We analyzed a sample of 138,374 women from 28 countries, 13 from Eastern and Southern Africa and 15 from West and Central Africa. DFPS ranged between 26.4% in Angola and 86.5% in Zimbabwe (Table 1).

**Table 1.** Demand for family planning satisfied by any contraceptive method (DFPS) in the 28 countries.

| Country | Survey<br>year | DFPS |  |
| --- | --- | --- | --- |
|  |  | % | N (unweighted) |
| West & Central Africa |  |  |  |
| Benin | 2017 | 32.4 | 5350 |
| Burkina Faso | 2021 | 68.6 | 6441 |
| Cameroon | 2018 | 45.6 | 3323 |
| Côte d'Ivoire | 2021 | 48.5 | 4034 |
| Gabon | 2019 | 42.6 | 2722 |
| Gambia | 2019 | 43.9 | 3381 |
| Ghana | 2022 | 60.8 | 5011 |
| Guinea | 2018 | 33.0 | 2510 |
| Liberia | 2019 | 42.7 | 2787 |
| Mali | 2018 | 42.0 | 3186 |
| Mauritania | 2019 | 31.5 | 4447 |
| Niger | 2021 | 45.5 | 2021 |
| Nigeria | 2018 | 46.9 | 10219 |
| Senegal | 2019 | 55.4 | 2788 |
| Sierra Leone | 2019 | 46.1 | 4403 |
| Eastern & Southern Africa |  |  |  |
| Angola | 2015 | 26.4 | 3849 |
| Burundi | 2016 | 49.0 | 5572 |
| Ethiopia | 2016 | 61.7 | 4894 |
| Kenya | 2022 | 81.7 | 6779 |
| Madagascar | 2021 | 77.3 | 7348 |
| Malawi | 2015 | 76.0 | 12434 |
| Mozambique | 2022 | 49.8 | 4532 |
| Rwanda | 2019 | 82.5 | 5662 |
| South Africa | 2016 | 78.6 | 1953 |
| Tanzania | 2022 | 64.2 | 5288 |
| Uganda | 2016 | 57.9 | 7556 |
| Zambia | 2018 | 71.6 | 5211 |
| Zimbabwe | 2015 | 86.5 | 4673 |

The two regions presented similar levels of empowerment within each age group and empowerment category. Additionally, within each empowerment and age subgroup, the mean SWPER scores were consistent across wealth quintiles and area of residence, with minimal variation observed.

### Demand for family planning satisfied according to women’s empowerment and age

Figure 1 shows the overall levels of DFPS according to women’s age and empowerment across the three SWPER domains. DFPS was higher in Eastern & Southern Africa than in West & Central Africa and clear empowerment-related trends were identified in both regions. Among women 20 years or older, DFPS increased with the level of empowerment in the three SWPER domains. For women aged 15 to 19, a consistent positive trend was identified only in the decision-making domain. Although DFPS was lower among adolescents with low empowerment in the domains of attitude to violence and social independence, a linear increase in coverage with the increase in the level of empowerment was not observed.

**Figure 1.**
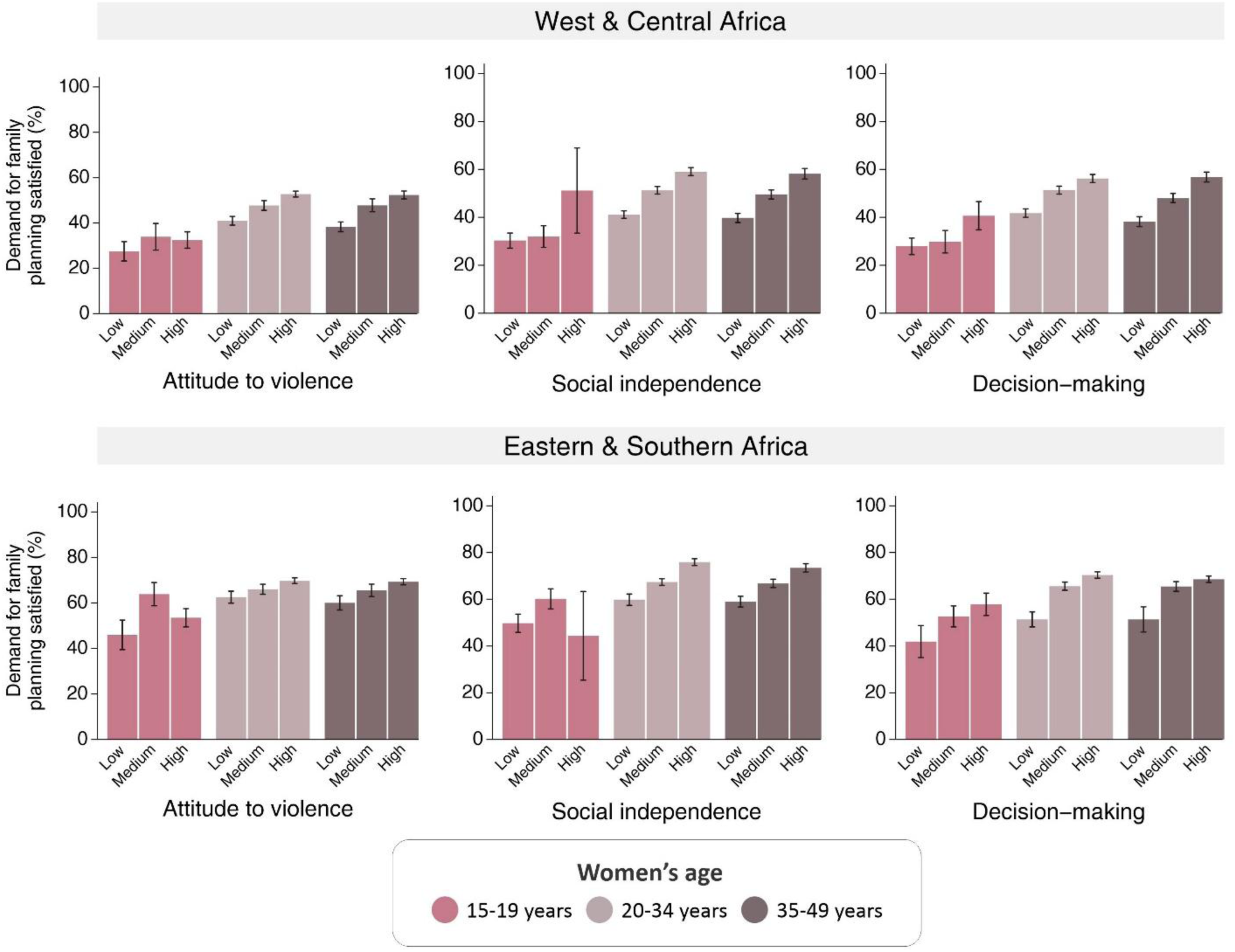
Pooled coverage of demand for family planning satisfied by any method according to region and women’s empowerment level.

In West & Central Africa, the most significant differences were identified in terms of social independence, where the DFPS among high-empowered women aged 15-19, 20-34, and 35-49 was 1.7, 1.4, and 1.5 times higher, respectively, than among the low-empowered. In the decision-making domain, adolescents with low and medium empowerment levels showed similar DFPS, but coverage was 50% higher among the high-empowered. For adult women, DFPS was 1.3 and 1.5 times higher among high-empowered women aged 20-34 and 35-49, respectively, than among low-empowered women. In the attitude to violence domain, DFPS among high-empowered women aged 15-19, 20-34, and 35-49 was 1.2, 1.3, and 1.4 times higher than among the low-empowered (Figure 1).

In Eastern & Southern Africa, the level of coverage was generally higher for all women, with slightly smaller differences across empowerment levels. However, important differences were identified in terms of decision-making, where the DFPS among high-empowered women aged 15-19 and 20-34 was 40% higher than among the low-empowered, and among women aged 35-49, it was 30% higher among the high-empowered (Figure 1).

### Method mix

The contraceptive method mix across the three domains of women’s empowerment and by region is presented in Figure 2. Regardless of empowerment level, the method mix in Eastern & Southern Africa was notably more skewed towards injectables compared to West & Central Africa. Implants accounted for 41.5% of contraceptive use in Eastern & Southern Africa and 23.3% in West & Central Africa. The most significant changes according to women’s empowerment were observed within the social independence domain. In Eastern & Southern Africa, the share of injectables was 33.7% lower among high-empowered women compared to the low-empowered (33.8% vs. 51.0%, respectively). This decrease was parallel with increases in the shares of condoms, fertility-awareness based methods, and pills, which were 3.1, 2.8, and 1.9 times higher among high-empowered women, respectively. In West & Central Africa, the largest variations were identified in the share of implants, which decreased from 30.5% among the low-empowered women to 21.0% among the high-empowered. The shares of condom and withdrawal increased significantly, from 3.7% to 10.9% and from 5.5% to 12.1%, respectively, between low- and high-empowered women. Similar trends were observed across the domains of attitude to violence and decision-making.

**Figure 2.**
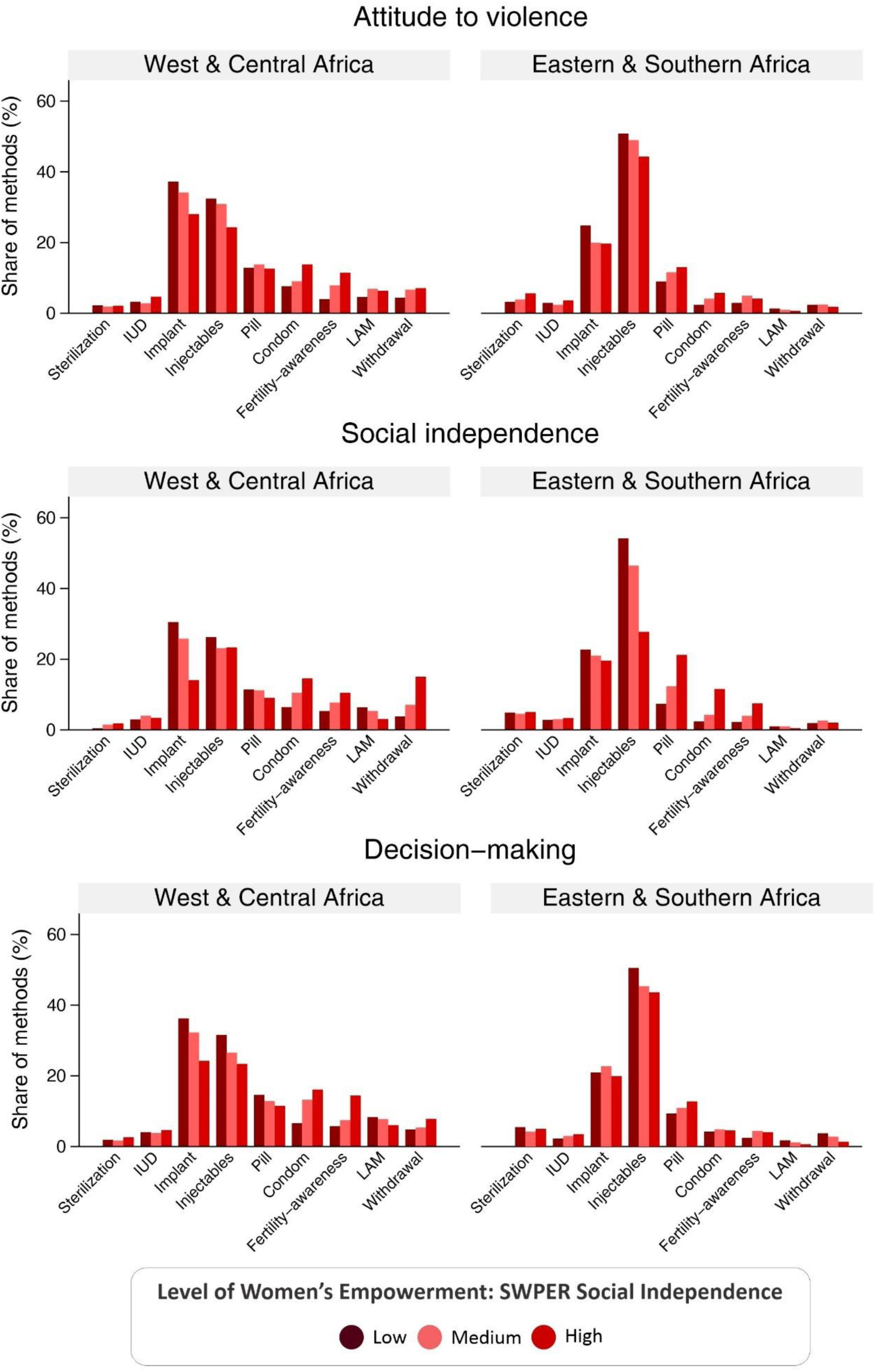
Pooled contraceptive method mix according to region and empowerment.

In terms of the intersectional differences in contraceptive method mix according to women’s empowerment and age, similar trends were also identified in the three SWPER domains, with more significant results according to women’s social independence than in decision-making and attitude to violence. Therefore, the results of these two domains are presented in the supplementary material, along with 95% confidence intervals for the three domains. The shares of each method by women’s age and empowerment within the social independence domain are presented in Figures 3 and 4.

**Figure 3.**
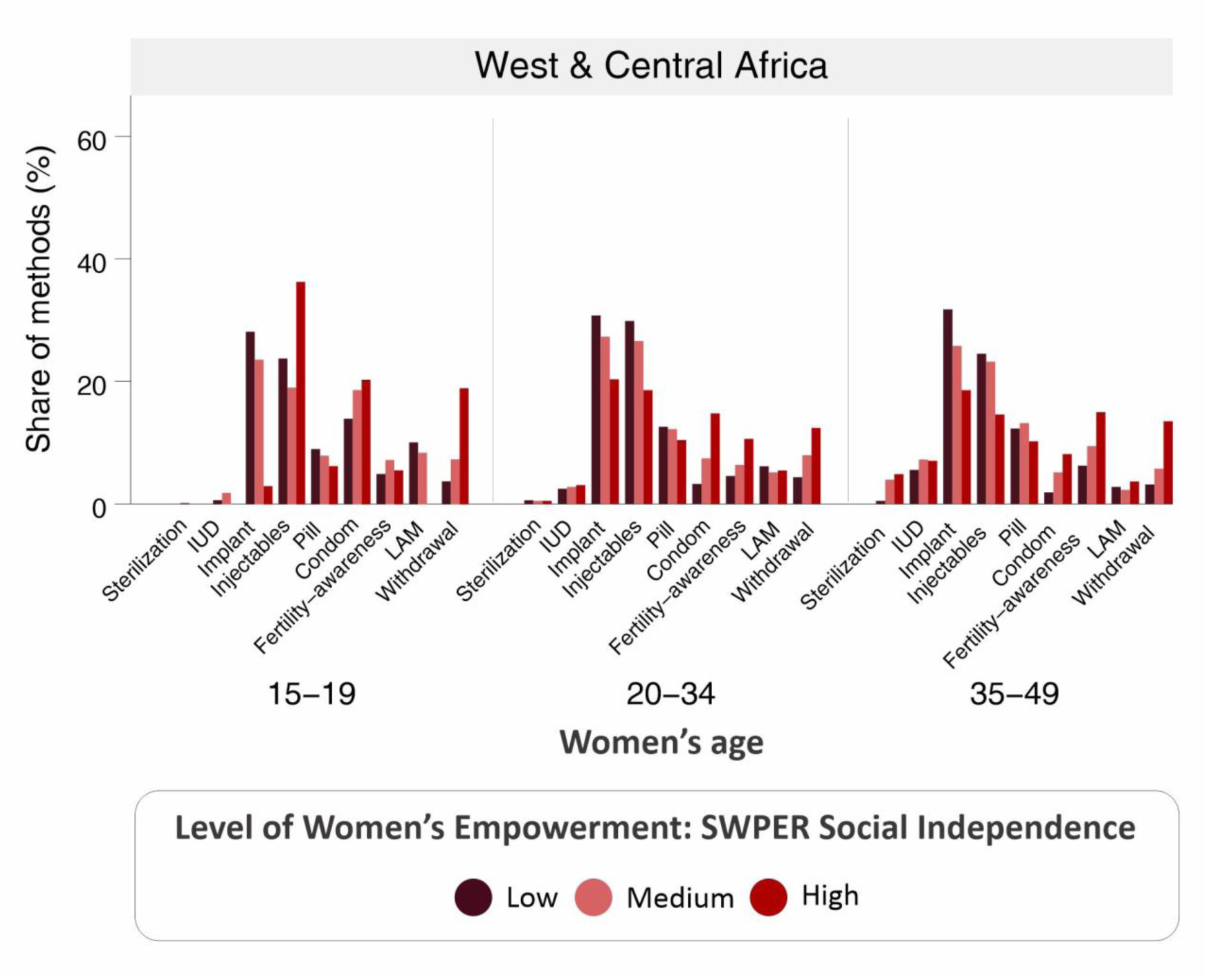
Pooled contraceptive method mix according to women’s age and empowerment level in the social independence domain (West & Central Africa).

**Figure 4.**
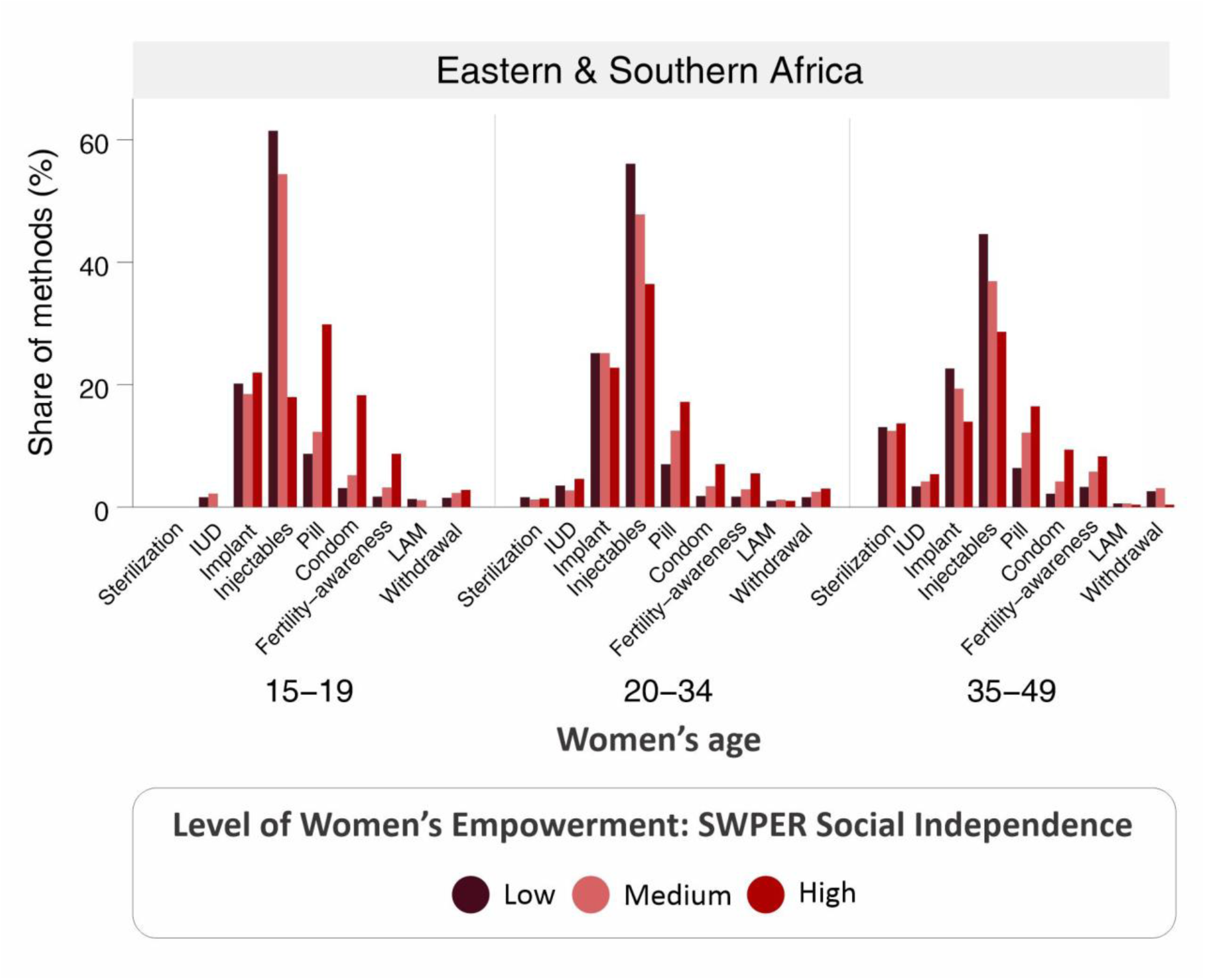
Pooled contraceptive method mix according to women’s age and empowerment level in the social independence domain (Eastern & Southern Africa).

The share of implants showed stronger reductions among women from West & Central Africa. Among those with high social independence, the share of implants dropped by 90%, 30%, and 40% for those aged 15-19, 20-34, and 35-49, respectively, compared to low-empowered women. In parallel, two methods—male condoms and fertility-awareness based methods—showed substantial increases with the increase in the level of empowerment. Among high-empowered adolescents, condom use increased by 50%, while among high-empowered adult women, it was over four times higher than among low-empowered women. Regarding the fertility-awareness based methods, its share was 2.3 and 2.4 times higher among high-empowered women aged 20-34 and 35-49, respectively. Among adolescents, no significant change was observed. Other contraceptive methods showed smaller and more variable changes (Figure 3).

In Eastern & Southern Africa, the largest reduction was identified in the share of injectables, which was reduced by 70%, 30%, and 40% among high-empowered women aged 15-19, 20-34, and 35-49, respectively. Decreases in the share of implants were also identified, especially among older women. Among the high-empowered women aged 35-49, the share of implants dropped by 40%. Conversely, there were consistent increases in the share of male condoms, pills, and fertility-awareness based methods across age groups. Among adolescents, the shares of condoms, pills, and fertility-awareness based methods were 3.4, 5.9, and 5.1 times higher, respectively, among high-empowered adolescents. In the 20-24 age group, these shares were 3.9, 2.5, and 3.2 times higher, while among women aged 35-49, they were 4.3, 2.6, and 2.5 times higher among high-empowered women compared to the low-empowered. Consistent increases were also identified in the share of IUDs among older women. It was 1.6 times higher among high-empowered women aged 35-49 in comparison to those who were low-empowered. No significant changes were observed in the share of other methods (Figure 4).

## Discussion

Our findings reveal increases in DFPS and a clear pattern of change in the contraceptive method mix according to women’s age and level of empowerment. In the three SWPER domains, we identified higher levels of DFPS and a more balanced method mix among high-empowered women, regardless of age. DFPS was higher in Eastern & Southern Africa than in West & Central Africa, with more consistent trends identified in terms of women’s decision-making in both regions. Regarding the contraceptive method mix, more significant changes were identified in the social independence domain. While the method mix was slightly different across regions, similar trends in terms of women’s age and empowerment were identified in both regions.

Lower levels of family planning coverage have been persistently identified in West & Central Africa (16,17). Although this low coverage has been historically linked to low use of modern contraceptives (20), our findings indicate that the method mix in West & Central Africa was predominantly represented by implants and injectables. The shares of condoms, LAM, and withdrawal were somewhat higher than in Eastern & Southern Africa, where the method mix was highly skewed towards injectables. Regardless of region, the shares of injectables and implants were significantly reduced with the increase in the level of empowerment. Reductions that happened in parallel with increases in the proportion of male condoms, pills, and fertility-awareness based methods.

Concentrating on a few methods has been used as a strategy to increase overall contraceptive use (2,4,7,8,21–23). This approach is simpler administratively and provides women with access to some form of contraception. However, there has been debate about whether family planning programs should prioritize expanding the acceptability and accessibility of less popular or newer methods or focus on increasing the availability of already widely accepted methods (23). The pathways for achieving broader contraceptive coverage and a method mix that aligns with women’s preferences, however, vary across different contexts. A sharp concentration of policies on only a few specific methods restricts choice and, therefore, is not aligned with the human rights-based approach to family planning and may not accommodate all women’s needs who may experience considerable side effects with available options. In sub-Saharan Africa, implants and injectables have been massively promoted as they can be easily provided through facility and community-based modalities, are less sensible to resupply issues as they last for more than only one reproductive cycle, and their efficacy is guaranteed as they are not dependent on users’ routine habits (22,23). Consequently, the introduction of injectables and implants in sub-Saharan Africa has been associated with significant increases in contraceptive use (20,24).

The emphasis on specific contraceptive methods within family planning programs has been influenced by various factors, including the cost and complexity of providing a diverse range of contraceptives, as well as the idea that certain methods are more effective in preventing unintended pregnancies (2). However, the effectiveness of each contraceptive method strongly depends on women’s ability to use them properly (25–27). Rather than unchangeable, this ability can be improved through family planning counseling, education, and women’s empowerment. When used correctly, most of the contraceptive methods have similar levels of efficacy (28). The availability of a wide range of contraceptive options is a central characteristic of high-quality family planning services and any condition that constrains a woman’s ability to make a full, free, and informed choice, such as coercion by programs or providers and inadequate contraceptive supplies, violates her rights and must be addressed (29). Therefore, all methods must be acknowledged as legitimate options, and healthcare providers must be adequately trained to offer them.

Among the three SWPER domains, the most strongly associated with DFPS was the decision-making, which considers the woman’s freedom to decide on visiting friends or relatives, on significant household purchases and on her own healthcare, while changes in the method mix were more significant in terms of women’s social independence. The social independence domain comprises aspects of women’s educational attainment, age at marriage and first child, access to mass media, and differences in age and education to the husband or cohabiting partner. The impact of women’s education on women’s sexual and reproductive health is among the most studied associations (16,30–33). More educated women tend to have more positive health-seeking attitudes and are better able to obtain, retain, and follow family planning counseling (32,33). Women’s age at marriage and age at first child, as well as age and educational differences to the husband, are important markers of family structure that have also been associated with family planning coverage (30). Earlier ages in these events and larger age differences between partners have been associated with more rigid social norms and lower levels of women’s agency (34). Other studies have also shown the effectiveness of access to mass media in increasing contraceptive use (30,32,33,35). More than just providing information, mass media channels, such as newspapers, magazines, and digital technologies, can increase the knowledge of women’s health and family planning options and combat myths about some family planning methods. As a result, it can build women’s self-efficacy and promote attitudes and behavioral changes (33,35). Our findings align with these associations, highlighting the stronger capability of more empowered women to navigate within the health system to choose the method that is best suited for them.

With age, women’s empowerment interacts in various ways. While empowerment often increases with age, influenced by factors such as labor-force participation and shifting roles within families and households, younger cohorts tend to be more empowered than older cohorts due to changing societal norms and structures (36). This intersection of age and empowerment affects contraceptive needs and preferences. By analyzing contraceptive method mix within each age group, we identified how empowerment at different life stages shapes the contraceptive method mix. Our findings indicate that high empowerment correlates with reduced reliance on implants and injectables across all ages, though specific shifts in method preference vary. Among adolescents and young adults, increased empowerment was associated with substantial rises in the shares of condom and fertility-awareness based methods. For older women, high empowerment was also linked to higher use of IUDs and sterilization.

Condoms and fertility-awareness based methods have important roles in such cases. Although our findings reveal a low share of condoms in the contraceptive method mix, substantial increases were identified with the increase in the level of women’s empowerment. This trend is consistent with the literature on gender norms surrounding condom use. A common barrier to condom use is the perception that it reduces men’s sexual pleasure (37). While all the hormonal contraceptives can also affect women’s sexual desire and pleasure (38,39), this side effect receives less attention, as gender norms implicitly or explicitly position sexual pleasure as a male privilege (37). Another common barrier to condom use among married couples is the women’s stigma of infidelity (40,41). Additionally, condoms are among the few contraceptive methods involving both partners, making partner cooperation essential. In consequence, low condom use has been partly attributed to women’s inability to negotiate its use with their partners (40).

Side effects and health concerns are the most common reasons for nonuse or discontinuation of hormonal contraceptives among women who need family planning services (42–46). While it is commonly assumed that women use non-modern contraceptives due to a lack of access or misconceptions regarding modern methods, these choices often reflect their genuine preferences, especially for those seeking more natural options (47). Fertility-awareness based methods have been used for centuries. However, they have significantly evolved over time. Whereas past techniques relied on the basic knowledge that conception is possible only on certain days following menstruation, modern approaches involve tracking multiple fertility biomarkers that reflect personal hormonal changes throughout a woman’s ovulation cycle—such as menstrual start dates, basal body temperature, cervical mucus, and sensations at the vulva (47,48). Unfortunately, national health surveys do not collect information on how effectively women are using these methods. Although incorrect knowledge of ovulation timing remains common in many African countries, younger, more educated, and women with internet access tend to have higher awareness (49–51). Additionally, the effectiveness of fertility-awareness methods also depends on couples’ sexual behavior during the fertile period – an aspect that is strongly related to women’s ability to negotiate sexual activity. Aspects that improve with women’s empowerment and align with our finding of higher use among highly empowered young women.

Women’s empowerment is a complex multidimensional construct (52–54). Beyond gender dynamics, it is linked to key contextual factors such as wealth and area of residence, both influencing and being influenced by them. While these interactions may partly shape the observed patterns, the standardized cut-offs of the SWPER Global help mitigate confounding by ensuring that women within each empowerment level have comparable levels of empowerment. Our findings from the sensitivity analysis further support this, showing that within each empowerment and age subgroup, the mean empowerment scores are consistent across urban and rural settings as well as wealth quintiles. On the other hand, the approach taken to generate the SWPER categories can result in small sample sizes for certain groups, which in this study was particularly evident among high-empowered adolescents. Although this is a considerable limitation, the stratification by women’s age remains valuable as it allows for the exploration of differences in contraceptive method mix by women’s empowerment across different life stages. Furthermore, the SWPER Global is limited to women who are married or in a union, excluding never-married women who may also have a demand for family planning services.

Unfortunately, surveys do not measure the method preferred by women, but rather the methods used. The lack of this information limits the interpretation of our findings. While there is evidence that availability and choice are closely related, with method preference following method availability (3), the method used is certainly influenced by other barriers such as proper knowledge of all contraceptives, costs associated with each method and opposition by others.

No ideal target level of use can be set for a specific method. However, considering the multiple circumstances women may be exposed to, their different desires, and the different side effects of each method, it is natural to ideally expect a more balanced method mix. Therefore, policymakers should consider the patterns of change in the method mix among high-empowered women in the development of their initiatives.

## Conclusion

Our findings contribute to a nuanced understanding of how empowerment, age, and the type of contraceptive method used intersect in ways that should be respected and supported within family planning policies. They highlight that women’s empowerment translates into diverse contraceptive preferences, sometimes counter to conventional family planning expectations and priorities regarding contraceptive choices. While national policies often emphasize certain methods based on financial, logistical, and effectiveness considerations, women may prioritize methods that offer greater personal control and fewer side effects. Given that the effectiveness of most contraceptive methods is closely linked to a woman’s ability to use them confidently and correctly, our findings underscore the importance of family planning counseling, comprehensive education, and targeted empowerment efforts. Such initiatives can enable women to make informed choices that align with their preferences and circumstances while ensuring that policies strike a balance between national priorities and the need to offer methods that address the diverse preferences of all women.

Recognizing that contraceptive preferences are shaped by an interplay of factors including relationship dynamics, personal autonomy, and exposure to family planning policies and programs, further research is needed to explore how these dimensions influence method choices and shifts in contraceptive use. Such research can guide the development of more inclusive and responsive family planning policies that respect and support women’s agency and reproductive health rights.

## Contributors

FH: conceptualization, formal analysis, interpretation of results, writing original draft; AB: conceptualization, supervision, and validation; YDW, CMF, JR, LA, RMK, TB, and AJDB critically revised the manuscript; All authors approved the final version of the manuscript. AB accessed and verified the data underlying this study.

## Data sharing statement

All data relevant to the study are included in the article or available as supplementary information. The data used in the analyses is publicly available, anonymized and geographically scrambled to ensure confidentiality. More information on DHS can be found at https://dhsprogram.com/, where survey datasets can be obtained.

## Data Availability

All data produced in the present work are contained in the manuscript.

## Acknowledgements

We thank Cintia Borges for her assistance in the graphs production and Kristin Bietsch for reviewing the article.

