## Supplementary material for "Women’s empowerment and life stage: assessing intersectional differences in contraceptive method mix in sub-Saharan Africa"

Supplementary Figure 1. Pooled contraceptive method mix according to region, women's age and empowerment level in the attitude to violence domain.

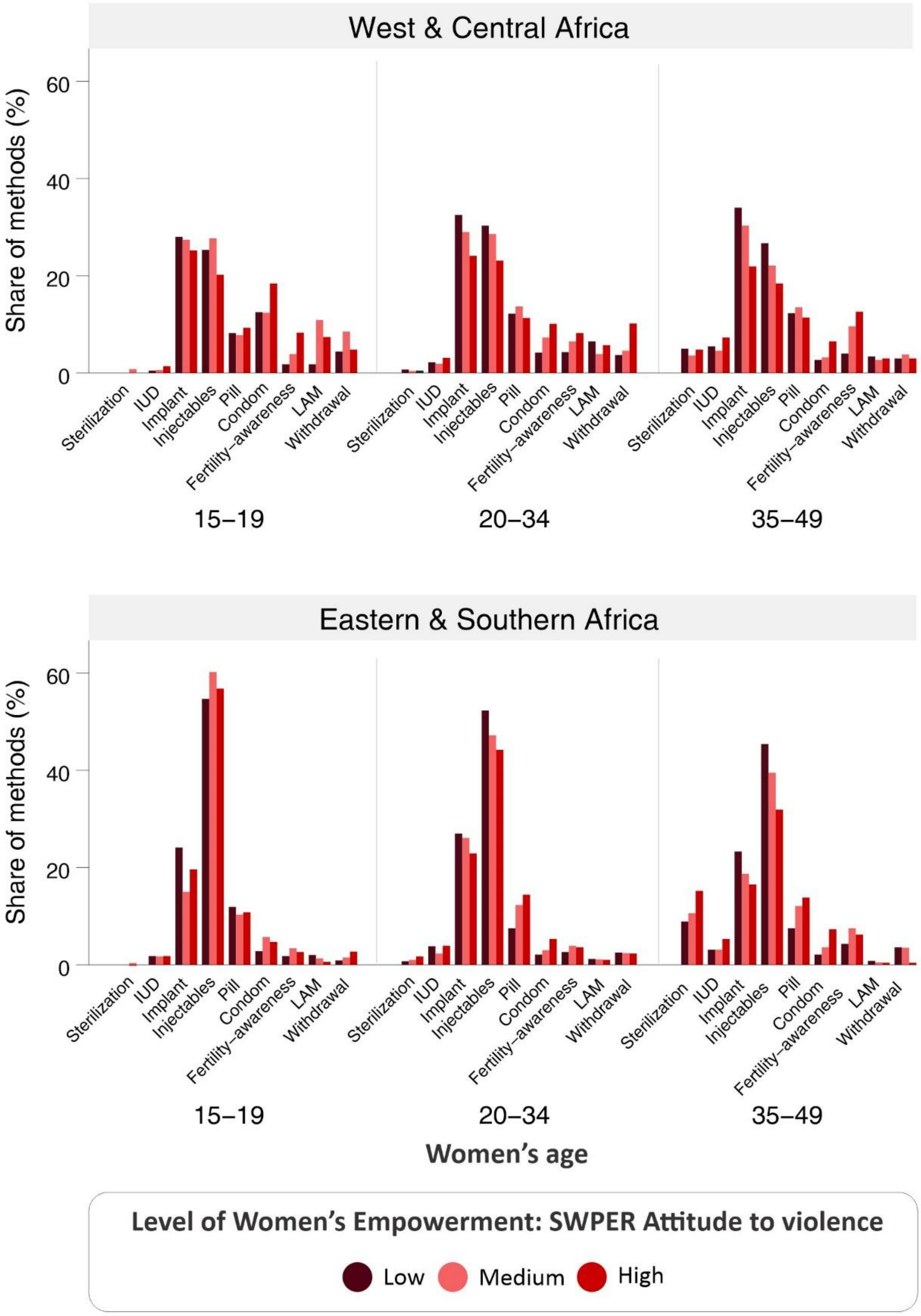

Supplementary Figure 2. Pooled contraceptive method mix according to region, women's age and empowerment level in the decision-making domain.

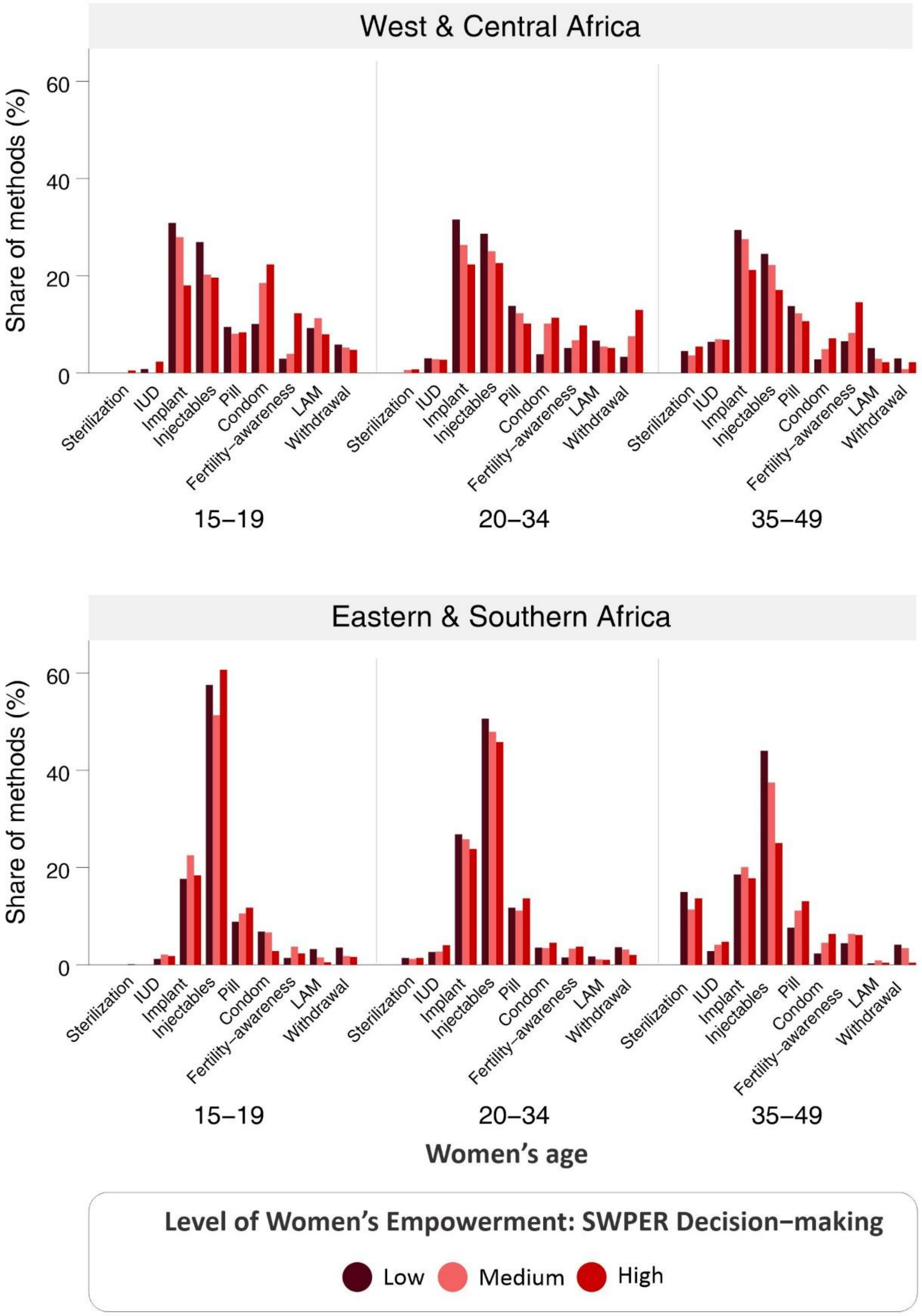

Supplementary Table 1. Mean women's empowerment scores (SWPER Global – social independence) by category of women's empowerment, age, and region according to women's household wealth and place of residence.

| Level of empowerment | Women's age | Wealth | | | | | Place of residence | |
|---|---|---|---|---|---|---|---|---|
| | | **Poorest** | **Poorer** | **Middle** | **Wealthier** | **Wealthiest** | **Urban** | **Rural** |
| | | Mean (SD) | Mean (SD) | Mean (SD) | Mean (SD) | Mean (SD) | Mean (SD) | Mean (SD) |
| **West & Central Africa** | | | | | | | | |
| | 15-19 | -1.15 (0.01) | -1.15 (0.01) | -1.13 (0.01) | -1.15 (0.02) | -1.00 (0.02) | -1.09 (0.02) | -1.14 (0.01) |
| Low | 20-34 | -1.15 (0.01) | -1.14 (0.01) | -1.12 (0.01) | -1.10 (0.00) | -1.06 (0.01) | -1.09 (0.01) | -1.14 (0.00) |
| | 35-49 | -1.14 (0.01) | -1.16 (0.01) | -1.15 (0.01) | -1.13 (0.01) | -1.09 (0.02) | -1.13 (0.01) | -1.15 (0.01) |
| | 15-19 | -0.31 (0.01) | -0.28 (0.01) | -0.24 (0.02) | -0.22 (0.01) | -0.22 (0.02) | -0.22 (0.01) | -0.27 (0.01) |
| Medium | 20-34 | -0.21 (0.00) | -0.20 (0.01) | -0.17 (0.01) | -0.14 (0.01) | -0.09 (0.01) | -0.13 (0.00) | -0.19 (0.00) |
| | 35-49 | -0.19 (0.01) | -0.18 (0.01) | -0.16 (0.01) | -0.14 (0.01) | -0.11 (0.01) | -0.13 (0.01) | -0.18 (0.00) |
| | 15-19 | 0.49 (0.03) | 0.47 (0.03) | 0.62 (0.08) | 0.52 (0.05) | 0.70 (0.09) | 0.67 (0.07) | 0.50 (0.03) |
| High | 20-34 | 0.74 (0.02) | 0.77 (0.01) | 0.87 (0.01) | 0.96 (0.01) | 1.21 (0.02) | 1.09 (0.01) | 0.90 (0.01) |
| | 35-49 | 0.94 (0.02) | 0.97 (0.02) | 1.1 (0.02) | 1.29 (0.02) | 1.59 (0.02) | 1.43 (0.02) | 1.15 (0.02) |
| **Eastern & Southern Africa** | | | | | | | | |
| | 15-19 | -1.00 (0.02) | -1.00 (0.02) | -0.96 (0.02) | -0.89 (0.02) | -0.94 (0.04) | -0.93 (0.02) | -0.99 (0.01) |
| Low | 20-34 | -1.05 (0.01) | -1.04 (0.01) | -1.04 (0.01) | -1.02 (0.01) | -1.03 (0.02) | -0.99 (0.01) | -1.05 (0.01) |
| | 35-49 | -1.10 (0.02) | -1.07 (0.01) | -1.07 (0.01) | -1.07 (0.01) | -1.05 (0.02) | -1.02 (0.01) | -1.08 (0.01) |
| | 15-19 | -0.27 (0.01) | -0.25 (0.02) | -0.25 (0.01) | -0.23 (0.01) | -0.21 (0.02) | -0.21 (0.01) | -0.26 (0.01) |
| Medium | 20-34 | -0.19 (0.01) | -0.18 (0.01) | -0.16 (0.01) | -0.13 (0.01) | -0.12 (0.01) | -0.11 (0.00) | -0.17 (0.00) |
| | 35-49 | -0.16 (0.01) | -0.16 (0.01) | -0.16 (0.01) | -0.16 (0.08) | -0.13 (0.01) | -0.12 (0.01) | -0.16 (0.00) |
| | 15-19 | 0.48 (0.04) | 0.65 (0.13) | 0.51 (0.05) | 0.46 (0.02) | 0.49 (0.04) | 0.46 (0.02) | 0.56 (0.05) |
| High | 20-34 | 0.77 (0.02) | 0.80 (0.02) | 0.87 (0.02) | 0.93 (0.01) | 1.13 (0.02) | 1.08 (0.01) | 0.84 (0.01) |
| | 35-49 | 1.01 (0.02) | 1.06 (0.02) | 1.15 (0.03) | 1.21 (0.02) | 1.43 (0.02) | 1.41 (0.02) | 1.06 (0.01) |

SD: standard error.

Supplementary Table 2. Pooled estimates of demand for family planning satisfied by any method (DFPS) and contraceptive method mix according to region, women's age and empowerment (attitude to violence).

| | **15-19** | | | **20-34** | | | **35-49** | | |
|---|---|---|---|---|---|---|---|---|---|
| | Low | Medium | High | Low | Medium | High | Low | Medium | High |
| **West & Central Africa** | | | | | | | | | |
| **Demand for family planning satisfied by any method** | 27.5 (23.2; 31.8) | 33.9 (28.0; 39.8) | 32.5 (28.9; 36.1) | 40.9 (39.0; 42.9) | 47.7 (45.6; 49.9) | 52.7 (51.4; 54.0) | 38.3 (36.1; 40.4) | 47.8 (44.9; 50.6) | 52.3 (50..6; 54.1) |
| **Method mix** | | | | | | | | | |
| Sterilization | 0.0 (0.0; 0.0) | 0.8 (0.0; 2.3) | 0.0 (0.0; 0.0) | 0.7 (0.2; 1.3) | 0.4 (0.1; 0.7) | 0.5 (0.3; 0.7) | 5.0 (3.5; 6.4) | 3.6 (2.1; 5.0) | 4.8 (4.0; 5.6) |
| IUD | 0.5 (0.0; 1.5) | 0.6 (0.0; 1.5) | 1.4 (0.0; 3.4) | 2.2 (1.6; 2.8) | 1.9 (1.3; 2.5) | 3.1 (2.5; 3.7) | 5.5 (4.4; 6.7) | 4.6 (3.2; 6.0) | 7.3 (6.4; 8.2) |
| Implants | 28.0 (20.6; 35.4) | 27.4 (19.3; 35.5) | 25.2 (19.9; 30.5) | 32.5 (29.7; 35.3) | 29.0 (26.5; 31.6) | 24.1 (22.6; 25.6) | 34.0 (31.1; 36.8) | 30.3 (26.7; 33.8) | 21.9 (20.2; 23.7) |
| Injectables | 25.3 (19.1; 31.5) | 27.7 (19.1; 36.3) | 20.2 (15.2; 25.2) | 30.3 (28.0; 32.7) | 28.6 (26.0; 31.2) | 23.1 (21.7; 24.5) | 26.7 (24.1; 29.3) | 22.1 (19.0; 25.1) | 18.4 (16.8; 20.1) |
| Pill | 8.2 (4.2; 12.1) | 7.8 (3.9; 11.8) | 9.3 (6.0; 12.5) | 12.2 (10.6; 13.8) | 13.7 (11.7; 15.8) | 11.3 (10.3; 12.3) | 12.3 (10.3; 14.3) | 13.5 (11.0; 15.8) | 11.4 (9.8; 12.9) |
| Male condom | 12.5 (5.8, 19.3) | 12.4 (6.5; 18.3) | 18.4 (13.2; 23.6) | 4.2 (3.0; 5.4) | 7.3 (5.8; 8.8) | 10.1 (9.0; 11.1) | 2.7 (1.4; 4.1) | 3.2 (2.1; 4.2) | 6.5 (5.4; 7.6) |
| Fertility-awareness based methods | 1.8 (0.1, 3.4) | 3.9 (0.8; 7.1) | 8.3 (4.9, 11.7) | 4.3 (3.3; 5.3) | 6.5 (5.1; 8.0) | 8.2 (7.3; 9.1) | 4.0 (3.0; 44.9) | 9.6 (7.4; 11.8) | 12.6 (11.3; 13.9) |
| Lactational amenorrhea | 1.8 (0.4; 3.1) | 10.9 (3.9; 17.8) | 7.4 (3.8; 10.9) | 6.5 (4.9; 17.6) | 3.9 (2.8; 5.0) | 5.7 (4.9; 6.6) | 3.4 (2.2; 4.6) | 2.7 (1.6; 3.7) | 3.0 (2.1; 3.9) |
| Withdrawal | 4.4 (1.2; 7.6) | 8.5 (2.3; 14.7) | 4.8 (1.0; 8.6) | 3.7 (2.6; 4.9) | 4.6 (3.3; 5.9) | 10.2 (8.7; 11.8) | 3.0 (1.7; 4.2) | 3.8 (2.2; 5.3) | 10.1 (8.4; 11.9) |
| Other modern | 0.0 (0.0; 0.0) | 0.0 (0.0; 0.0) | 0.1 (0.0; 0.2) | 0.3 (0.0; 0.5) | 0.2 (0.0; 0.5) | 0.2 (0.0; 0.3) | 0.2 (0.0; 0.5) | 0.7 (0.0; 1.5) | 0.1 (0.0; 0.2) |
| Other non-modern | 0.0 (0.0; 0.0) | 0.0 (0.0; 0.0) | 0.0 (0.0; 0.0) | 0.2 (0.0; 0.7) | 0.2 (0.0; 0.4) | 0.1 (0.0; 0.2) | 0.2 (0.0; 0.5) | 0.4 (0.0; 1.1) | 0.1 (0.0; 0.2) |
| **Eastern & Southern Africa** | | | | | | | | | |
| **Demand for family planning satisfied by any method** | 46.0 (39.5; 52.4) | 63.9 (58.8; 68.9) | 53.5 (49.5; 57.5) | 62.5 (59.9; 65.1) | 66.0 (63.8; 68.2) | 59.8 (68.5; 71.0) | 60.0 (56.9; 63.1) | 65.5 (62.8; 68.2) | 69.3 (68.0; 70.7) |
| **Method mix** | | | | | | | | | |
| Sterilization | 0.0 (0.0; 0.0) | 0.1 (0.0; 0.2) | 0.0 (0.0; 0.0) | 0.7 (0.3; 1.0) | 1.0 (0.6; 1.5) | 1.7 (1.4; 2.1) | 8.9 (7.5; 10.4) | 10.6 (9.2; 12.1) | 15.2 (14.0; 16.5) |
| IUD | 1.8 (0.0; 4.9) | 1.7 (0.0; 4.4) | 1.8 (0.7; 3.0) | 3.8 (2.4; 5.2) | 2.3 (1.6; 2.9) | 3.9 (3.3; 4.5) | 3.1 (1.8; 4.30 | 3.1 (2.2; 4.0) | 5.3 (4.5; 6.0) |
| Implants | 24.1 (17.2; 31.0) | 15.1 (9.9; 20.3) | 19.6 (15.9; 23.3) | 27.0 (24.6; 29.5) | 26.1 (24.2; 28.0) | 22.9 (21.9; 23.9) | 23.3 (20.7; 25.8) | 18.7 (16.7; 20.7) | 16.5 (15.4; 17.6) |
| Injectables | 54.7 (46.1; 63.2) | 60.2 (53.0; 67.3) | 56.8 (52.2; 61.4) | 52.3 (49.4; 55.2) | 47.2 (44.8; 49.7) | 44.2 (42.7; 45.6) | 45.4 (42.1; 48.8) | 39.5 (36.3; 42.6) | 31.9 (30.3; 33.4) |
| Pill | 11.9 (6.8; 17.0) | 10.3 (7.3; 13.3) | 10.8 (7.1; 14.5) | 7.5 (6.4; 8.6) | 12.3 (10.9; 13.6) | 14.4 (13.3; 15.4) | 7.5 (6.0; 8.9) | 12.1 (10.6; 13.6) | 13.8 (12.8; 14.9) |
| Male condom | 2.8 (1.4, 4.2) | 5.7 (1.6; 2.6) | 4.7 (3.0; 6.4) | 2.1 (1.6; 2.5) | 3.0 (2.3; 3.7) | 5.3 (4.7; 5.9) | 2.1 (1.6; 2.6) | 3.6 (2.7; 4.6) | 7.3 (6.5; 8.2) |
| Fertility-awareness based methods | 1.8 (0.4; 3.1) | 3.4 (0.6; 6.3) | 2.6 (1.1; 4.1) | 2.6 (2.0; 3.2) | 3.9 (3.3; 4.6) | 3.6 (3.2; 4.0) | 4.3 (3.4; 5.2) | 7.5 (6.1; 8.8) | 6.2 (5.6; 6.8) |
| Lactational amenorrhea | 2.0 (0.0; 3.4) | 1.3 (0.0, 3.4) | 0.6 (0.1; 1.2) | 1.2 (0.8; 1.5) | 1.1 (0.8; 1.5) | 1.0 (0.8; 1.2) | 0.8 (0.4; 1.2) | 0.5 (0.2; 0.7) | 0.4 (0.3; 0.5) |
| Withdrawal | 0.9 (0.1; 1.7) | 1.5 (0.5; 2.4) | 2.7 (1.4; 4.1) | 2.5 (1.9; 3.2) | 2.4 (1.9; 2.9) | 2.3 (2.0; 2.7) | 3.6 (2.8; 4.3) | 3.5 (2.6; 4.5) | 2.4 (2.0; 2.8) |
| Other modern | 0.0 (0.0; 0.0) | 0.2 (0.0; 0.5) | 0.0 (0.0; 0.1) | 0.0 (0.0; 0.0) | 0.0 (0.0; 0.0) | 0.2 (0.0; 0.3) | 0.1 (0.0; 0.1) | 0.2 (0.0; 0.3) | 0.2 (0.0; 0.3) |
| Other non-modern | 0.0 (0.0; 0.0) | 0.0 (0.0; 0.0) | 0.0 (0.0; 0.1) | 0.0 (0.0; 0.0) | 0.0 (0.0; 0.0) | 0.1 (0.0; 0.2) | 0.0 (0.0; 0.0) | 0.1 (0.0; 0.3) | 0.1 (0.0; 0.3) |

Supplementary Table 3. Pooled estimates of demand for family planning satisfied by any method (DFPS) and contraceptive method mix according to region, women's age and empowerment (social independence).

| | **15-19** | | | **20-34** | | | **35-49** | | |
|---|---|---|---|---|---|---|---|---|---|
| | Low | Medium | High | Low | Medium | High | Low | Medium | High |
| **West & Central Africa** | | | | | | | | | |
| **Demand for family planning satisfied by any method** | 30.3 (27.2; 33.5) | 32.0 (27.5; 36.5) | 51.2 (33.4; 68.9) | 41.2 (39.6; 42.7) | 51.3 (49.7; 52.9) | 59.1 (57.4; 60.7) | 39.8 (37.8; 41.7) | 49.5 (47.6; 51.4) | 58.2 (56.0; 60.4) |
| **Method mix** | | | | | | | | | |
| Sterilization | 0.2 (0.0; 0.6) | 0.0 (0.0; 0.0) | 0.0 (0.0; 0.0) | 0.6 (0.3; 0.9) | 0.5 (0.2; 0.7) | 0.5 (0.2; 0.8) | 0.5 (0.4; 0.6) | 4.0 (2.8; 5.2) | 4.9 (0.4; 0.6) |
| IUD | 0.7 (0.1; 1.3) | 1.9 (0.0; 5.3) | 0.0 (0.0; 0.0) | 2.5 (2.0; 3.1) | 2.8 (0.0; 3.6) | 3.1 (2.2; 4.1) | 5.6 (4.6; 6.7) | 7.3 (6.0; 8.6) | 7.1 (5.9; 8.4) |
| Implants | 28.4 (23.5; 33.4) | 23.8 (17.4; 30.2) | 3.9 (0.0; 0.9) | 31.0 (29.1; 32.9) | 27.5 (25.6; 29.4) | 20.5 (18.5; 22.4) | 32.0 (29.3; 34.6) | 26.0 (23.9; 28.3) | 18.7 (16.7; 20.8) |
| Injectables | 24.0 (19.3; 28.7) | 19.2 (13.4; 24.9) | 36.6 (11.2; 62.0) | 30.1 (28.3; 32.0) | 26.8 (25.0; 28.6) | 18.7 (16.8; 20.5) | 24.7 (22.4; 26.9) | 23.4 (21.3; 25.6) | 14.7 (12.9; 16.6) |
| Pill | 9.1 (6.4; 11.8) | 8.0 (4.3; 11.7) | 6.3 (0.0; 11.7) | 12.7 (11.3; 14.1) | 12.3 (10.9; 13.7) | 10.5 (9.2; 11.7) | 12.4 (10.8; 14.0) | 13.3 (10.8; 15.7) | 10.3 (8.7; 11.9) |
| Male condom | 14.1 (9.7; 18.5) | 18.8 (11.6; 26.0) | 20.5 (0.0; 41.9) | 3.3 (2.5; 2.6) | 7.5 (6.1; 8.8) | 14.9 (13.2; 16.5) | 1.9 (1.2; 2.6) | 5.2 (3.8; 6.5) | 8.2 (6.6; 9.9) |
| Fertility-awareness based methods | 5.0 (2.8; 7.2) | 7.3 (2.9; 11.8) | 5.6 (0.0; 12.3) | 4.6 (3.4; 5.4) | 6.4 (5.5; 7.3) | 10.7 (9.2; 12.3) | 6.3 (4.3; 8.3) | 9.5 (7.9; 11.1) | 15.1 (13.1; 17.0) |
| Lactational amenorrhea | 10.2 (6.7; 13.6) | 8.5 (4.0; 13.0) | 0.0 (0.0; 0.0) | 6.2 (5.0; 7.4) | 5.2 (4.1; 6.2) | 5.5 (4.3; 6.6) | 2.8 (2.0; 3.6) | 2.3 (1.4; 3.1) | 3.7 (2.2; 5.2) |
| Withdrawal | 3.8 (0.9; 6.8) | 7.4 (2.3; 12.5) | 19.1 (0.0; 41.4) | 4.4 (2.7; 6.0) | 8.0 (2.3; 12.5) | 12.5 (10.7; 14.4) | 3.2 (2.3; 4.1) | 5.8 (4.3; 7.2) | 13.6 (11.2; 16.0) |
| Other modern | 0.0 (0.0; 0.0) | 0.1 (0.0; 0.4) | 0.0 (0.0; 0.0) | 0.0 (0.0; 0.1) | 0.3 (0.0; 0.4) | 0.2 (0.0; 0.4) | 0.4 (0.0; 7.4) | 0.0 (0.0; 0.2) | 0.0 (0.0; 0.2) |
| Other non-modern | 0.0 (0.0; 0.0) | 0.0 (0.0; 0.0) | 0.0 (0.0; 0.0) | 0.0 (0.0; 0.1) | 0.2 (0.0; 0.4) | 0.2 (0.0; 0.3) | 0.3 (0.0; 0.6) | 0.1 (0.0; 0.2) | 0.0 (0.0; 0.2) |
| **Eastern & Southern Africa** | | | | | | | | | |
| **Demand for family planning satisfied by any method** | 49.7 (45.8; 53.6) | 60.2 (55.9; 64.5) | 44.4 (25.4; 63.3) | 59.8 (56.7; 61.3) | 67.4 (65.9; 68.8) | 75.9 (74.4; 77.4) | 59.0 (56.7; 61.3) | 66.8 (65.0; 68.6) | 73.4 (71.7; 75.2) |
| **Method mix** | | | | | | | | | |
| Sterilization | 0.0 (0.0; 0.1) | 0.0 (0.0; 0.0) | 0.0 (0.0; 0.0) | 1.6 (1.2; 2.0) | 1.2 (0.9; 1.5) | 1.4 (0.8; 1.9) | 13.1 (11.7; 14.5) | 12.5 (11.3; 13.7) | 13.7 (12.0; 15.3) |
| IUD | 1.6 (0.0; 3.2) | 2.2 (0.2; 4.1) | 0.0 (0.0; 0.0) | 3.5 (2.4; 4.6) | 2.7 (2.1; 3.3) | 4.6 (3.9; 5.3) | 3.4 (2.4; 4.4) | 4.2 (3.3; 5.0) | 5.4 (4.5; 6.3) |
| Implants | 20.2 (15.8; 24.5) | 18.5 (14.2; 22.8) | 22.0 (7.4; 36.5) | 25.2 (23.3; 27.2) | 25.2 (23.9; 26.7) | 22.8 (21.4; 24.2) | 22.7 (20.5; 25.0) | 19.4 (18.0; 20.8) | 14.0 (12.8; 15.2) |
| Injectables | 61.6 (56.1; 67.1) | 54.5 (49.0; 60.0) | 18.3 (6.3; 30.4) | 56.2 (53.8; 58.7) | 47.9 (46.2; 49.5) | 36.5 (34.7; 38.4) | 44.7 (41.9; 47.5) | 37.0 (34.9; 39.1) | 28.7 (26.7; 30.7) |
| Pill | 8.7 (5.5; 12.0) | 12.3 (9.2; 15.4) | 29.9 (6.6; 53.2) | 7.0 (6.1; 7.9) | 12.5 (11.4; 13.5) | 17.2 (15.8; 18.6) | 6.4 (5.4; 7.3) | 12.2 (11.0; 13.4) | 16.5 (15.1; 18.0) |
| Male condom | 3.1 (2.0; 4.1) | 5.2 (3.4; 7.0) | 18.3 (1.1; 35.5) | 1.8 (1.5; 2.2) | 3.4 (2.9; 3.9) | 7.01 (8.1; 10.7) | 2.2 (1.7; 2.7) | 4.2 (3.5; 4.9) | 9.4 (8.1; 10.7) |
| Fertility-awareness based methods | 1.7 (1.0; 2.5) | 3.2 (1.0; 5.5) | 8.7 (0.0; 19.0) | 1.7 (1.3; 2.1) | 2.9 (2.4; 3.3) | 5.5 (4.8; 6.2) | 3.3 (2.6; 4.1) | 5.8 (5.0; 6.5) | 8.3 (7.3; 9.3) |
| Lactational amenorrhea | 1.3 (1.1; 2.5) | 1.1 (0.0; 2.3) | 0.0 (0.0; 0.0) | 1.0 (0.6; 1.3) | 1.2 (0.9; 1.5) | 1.0 (0.8; 1.3) | 0.6 (0.3; 1.0) | 0.6 (0.4; 0.8) | 0.4 (0.2; 0.5) |
| Withdrawal | 1.5 (0.6; 2.5) | 2.3 (1.2 ; 3.5) | 2.8 (0.0; 6.2) | 1.6 (1.2; 1.9) | 2.5 (2.0; 3.0) | 3.0 (2.5; 3.5) | 2.6 (2.0; 3.2) | 3.1 (2.5; 3.6) | 2.8 (2.3; 3.3) |
| Other modern | 0.0 (0.0; 0.0) | 0.2 (0.0; 0.4) | 0.0 (0.0; 0.0) | 0.0 (0.0; 0.0) | 0.0 (0.0; 0.2) | 0.2 (0.0; 0.3) | 0.1 (0.0; 0.2) | 0.0 (0.0; 0.0) | 0.3 (0.0; 0.5) |
| Other non-modern | 0.0 (0.0; 0.0) | 0.0 (0.0; 0.0) | 0.0 (0.0; 0.0) | 0.0 (0.0; 0.0) | 0.0 (0.0; 0.2) | 0.0 (0.0 (0.0; 0.1) | 0.0 (0.0; 0.0) | 0.0 (0.0; 0.0) | 0.2 (0.0; 0.4) |

Supplementary Table 4. Pooled estimates of demand for family planning satisfied by any method (DFPS) and contraceptive method mix according to region, women's age and empowerment (decision-making).

| | **15-19** | | | **20-34** | | | **35-49** | | |
|---|---|---|---|---|---|---|---|---|---|
| | Low | Medium | High | Low | Medium | High | Low | Medium | High |
| **West & Central Africa** | | | | | | | | | |
| **Demand for family planning satisfied by any method** | 27.9 (24.5; 31.3) | 29.8 (25.2; 34.5) | 40.7 (34.8; 46.7) | 41.8 (40.0; 43.5) | 51.4 (49.7; 53.0) | 56.2 (54.5; 57.9) | 38.2 (36.2; 40.2) | 48.1 (46.1; 50.0) | 56.8 (54.8; 58.9) |
| **Method mix** | | | | | | | | | |
| Sterilization | 0.0 (0.0; 0.0) | 0.0 (0.0; 0.0) | 0.5 (0.0; 1.5) | 0.3 (0.0; 0.5) | 0.6 (0.3; 0.9) | 0.7 (0.3; 0.9) | 4.5 (3.2; 5.8) | 3.6 (2.7; 4.4) | 5.4 (4.4; 6.4) |
| IUD | 0.8 (0.0; 1.6) | 0.0 (0.0; 0.0) | 2.3 (0.0; 6.1) | 3.0 (2.3; 3.7) | 2.8 (2.1; 3.6) | 2.7 (1.9; 3.5) | 6.4 (5.1; 7.7) | 6.9 (5.5; 8.2) | 6.8 (5.7; 7.9) |
| Implants | 30.7 (24.5; 36.8) | 27.8 (20.5; 35.0) | 17.9 (11.3; 24.4) | 31.4 (29.4; 33.5) | 26.2 (24.3; 28.1) | 22.2 (24.3; 28.1) | 29.3 (26.7; 31.9) | 27.4 (24.9; 29.9) | 21.1 (19.0; 23.1) |
| Injectables | 26.8 (21.0; 32.7) | 20.1 (14.2; 25.9) | 19.5 (12.9; 26.1) | 28.5 (26.6; 30.4) | 24.9 (22.9; 27.0) | 22.5 (20.7; 24.3) | 24.4 (19.9; 24.3) | 22.1 (19.9; 24.3) | 17.0; (15.1; 19.0) |
| Pill | 9.4 (6.0; 12.8) | 8.0 (4.0; 16.1) | 8.3 (4.4; 12.1) | 13.7 (12.2; 15.2) | 12.2 (10.8; 13.5) | 10.1 (8.9; 11.3) | 13.7 (11.9; 15.6) | 12.2 (10.5; 13.9) | 10.6 (8.5; 12.8) |
| Male condom | 10.0 (5.7; 14.4) | 18.4 (11.7; 25.1) | 22.2 (13.7; 30.8) | 3.8 (3.0; 4.7) | 10.1 (8.7; 11.6) | 11.3 (10.0; 12.6) | 2.8 (1.7; 3.9) | 4.9 (3.6; 6.2) | 7.1 (5.7; 8.5) |
| Fertility-awareness based methods | 2.9 (1.2; 4.5) | 3.9 (1.0; 6.9) | 12.2 (6.4; 17.9) | 5.1 (4.2; 5.9) | 6.7 (5.3; 8.1) | 9.7 (8.4; 10.7) | 6.5 (5.1; 7.8) | 8.2 (6.7; 9.6) | 14.5 (12.8; 16.2) |
| Lactational amenorrhea | 9.2 (5.3; 13.0) | 11.2 (6.3; 16.1) | 7.9 (2.4; 13.4) | 6.6 (5.2; 7.9) | 5.4 (4.2; 6.6) | 5.1 (4.1; 6.0) | 5.1 (2.2; 8.1) | 2.9 (2.1; 3.8) | 2.2 (1.6; 2.9) |
| Withdrawal | 5.8 (1.3; 10.3) | 5.2 (1.1; 9.2) | 4.7 (0.3; 9.2) | 3.3 (2.4; 4.2) | 7.5 (6.1; 9.0) | 12.9 (10.5; 15.3) | 3.0 (2.0; 4.0) | 0.8 (5.8; 9.3) | 11.1 (9.1; 13.2) |
| Other modern | 0.0 (0.0; 0.0) | 0.2 (0.0; 0.5) | 0.0 (0.0; 0.0) | 0.1 (0.0; 0.3) | 0.3 (0.1; 0.5) | 0.2 (0.0; 0.3) | 0.2 (0.0; 0.4) | 0.3 (0.0; 0.6) | 0.1 (0.0; 0.2) |
| Other non-modern | 0.0 (0.0; 0.0) | 0.0 (0.0; 0.0) | 0.0 (0.0; 0.0) | 0.1 (0.0; 0.3) | 0.2 (0.0; 0.4) | 0.1 (0.0; 0.3) | 0.3 (0.0; 0.6) | 0.3 (0.0; 0.6) | 0.1 (0.0; 0.2) |
| **Eastern & Southern Africa** | | | | | | | | | |
| **Demand for family planning satisfied by any method** | 41.9 (35.1; 48.7) | 52.7 (48.2; 57.2) | 57.8 (53.1; 62.6) | 51.4 (48.2; 54.6) | 65.6 (63.9; 67.4) | 70.4 (69.0; 71.2) | 51.4 (46.0; 56.8) | 65.5 (63.4; 67.6) | 68.6 (67.2; 70.0) |
| **Method mix** | | | | | | | | | |
| Sterilization | 0.1 (0.0; 0.3) | 0.0 (0.0; 0.0) | 0.0 (0.0; 0.0) | 1.4 (0.8; 2.0) | 1.2 (0.9; 1.5) | 1.4 (1.1; 1.7) | 14.9 (12.0; 17.8) | 11.3 (10.1; 12.6) | 13.6 (12.4; 14.7) |
| IUD | 1.2 (0.0; 2.9) | 2.1 (0.0; 4.7) | 1.8 (0.0; 3.3) | 2.6 (1.2; 4.0) | 2.7 (2.1; 3.3) | 4.0 (3.3; 4.7) | 2.8 (1.2; 4.4) | 4.1 (2.9; 5.4) | 4.7 (4.0; 5.3) |
| Implants | 17.6 (10.2; 25.0) | 22.4 (17.4; 27.4) | 18.3 (14.3; 22.4) | 26.7 (23.6; 29.8) | 25.7 (24.1; 27.2) | 23.7 (22.6; 24.8) | 18.5 (15.0; 22.0) | 20.0 (18.3; 21.2) | 17.7 (16.5; 18.8) |
| Injectables | 57.3 (46.7; 67.8) | 51.1 (45.2; 57.0) | 60.4 (55.4; 65.4) | 50.4 (46.6; 54.2) | 47.7 (45.9; 49.5) | 45.6 (44.0; 47.2) | 43.8 (38.0; 49.5) | 37.3 (34.9; 39.6) | 24.9 (33.3; 36.6) |
| Pill | 8.8 (2.3; 15.3) | 10.5 (7.9; 13.2) | 11.7 (8.1; 15.3) | 11.7 (8.1; 15.3) | 11.1 (10.2; 12.1) | 13.6 (12.6; 14.6) | 7.6 (5.4; 9.8) | 11.1 (9.8; 12.4) | 13.0 (12.0; 14.0) |
| Male condom | 6.8 (2.2; 11.5) | 6.6 (4.3; 9.0) | 2.8 (1.8; 3.8) | 3.5 (2.4; 4.6) | 3.4 (3.0; 4.1) | 4.5 (3.9; 5.1) | 2.3 (3.6; 5.3) | 4.5 (3.6; 5.3) | 6.3 (5.5; 7.1) |
| Fertility-awareness based methods | 1.4 (0.0; 3.3) | 3.7 (1.0; 6.4) | 2.3 (1.0; 3.5) | 1.5 (1.0; 2.1) | 3.3 (2.8; 3.9) | 3.7 (3.3; 4.1) | 4.4 (5.2; 7.3) | 6.3 (5.2; 7.4) | 6.1 (5.5; 6.7) |
| Lactational amenorrhea | 3.2 (0.0; 7.0) | 1.5 (0.0; 3.3) | 0.5 (0.1; 0.9) | 1.7 (1.0; 2.5) | 1.1 (0.8; 1.2) | 1.0 (0.8; 1.2) | 0.3 (0.5; 1.3) | 0.9 (0.5; 1.3) | 0.4 (0.3; 0.5) |
| Withdrawal | 3.5 (0.7; 6.2) | 1.8 (0.8; 2.8) | 1.6 (0.7; 2.6) | 3.6 (2.5; 4.6) | 3.1 (2.5; 3.6) | 2.0 (1.7; 2.3) | 4.1 (2.6; 5.6) | 3.4 (2.7; 4.0) | 2.6 (2.2; 2.9) |
| Other modern | 0.0 (0.0; 0.0) | 0.0 (0.0; 0.0) | 0.1 (0.0; 0.3) | 0.3 (0.0; 0.8) | 0.1 (0.0; 0.2) | 0.1 (0.0; 0.1) | 0.0 (0.0; 0.0) | 0.1 (0.0; 0.1) | 0.2 (0.1; 0.3) |
| Other non-modern | 0.0 (0.0; 0.0) | 0.0 (0.0; 0.1) | 0.0 (0.0; 0.0) | 0.0 (0.0; 0.1) | 0.1 (0.0; 0.2) | 0.0 (0.0; 0.0) | 0.0 (0.0; 0.0) | 0.0 (0.0; 0.1) | 0.1 (0.0; 0.2) |
